# Deep sequencing of DNA from urine of kidney allograft recipients to estimate the donor-specific DNA fraction

**DOI:** 10.1101/2020.12.13.20248118

**Authors:** Aziz Belkadi, Gaurav Thareja, Darshana Dadhania, John R. Lee, Thangamani Muthukumar, Catherine Snopkowski, Carol Li, Anna Halama, Sara Abdelkader, Yasmin Mahmoud, Joel Malek, Manikkam Suthanthiran, Karsten Suhre

## Abstract

Renal transplantation is the method of choice for patients with end stage kidney failure. But transplanted allograft could be affected by viral and bacterial infections and immune rejections. The standard test for the diagnosis of acute pathologies in kidney transplants is the renal biopsy. However, noninvasive tests would be desirable. Various methods using different techniques have been developed by the transplantation community. But these methods expect improvements. We present here a cost-effective method based on estimating donor-specific DNA fraction in recipient urine based on sequencing of recipient urine DNA only. We hypothesized that in the no-pathology stage, the largest tissue types present in recipient urine are donor kidney cells and in case of rejection, a larger number of recipient immune cells would be observed. Extensive in-silico simulation was used to tune the sequencing parameters: number of variants and depth of coverage. Sequencing of DNA mixture from 2 healthy individuals showed the method high prediction accuracy (maximum error < 0.04). We then demonstrated the insignificant impact of familial relationship and ethnicity using an in-house and public database. Lastly, we performed recipient deep urine DNA sequencing in 32 samples representing two pathology groups: acute rejection (AR, 12 samples) and acute tubular injury (ATI, 11 samples) and 9 samples with no pathology. We found a significant association between the donor-specific DNA fraction in the two pathology groups compared to no pathology (P = 0.0064 for AR and P = 0.026 for ATI). We conclude that deep DNA sequencing of recipient urine offers a noninvasive means of diagnosing and prognosticating acute pathologies in the human kidney allograft.

## Introduction

In 1933 surgeon Yurii Voronoy from Ukraine achieved the first human kidney transplantation [1]. Kidney transplantation is the final treatment option for patients with end-stage renal failure after dialysis. Today, renal transplantation plays an important role in clinical medicine and has become a relatively safe intervention. However, various pathologies can still affect the transplanted organ, including infections, disease recurrence and immune rejections. These rejections can be related to a range of donor- and recipient-specific factor risks [2,3]. Acute renal rejection affects 10 to 20% of transplants within three months after transplantation and chronic rejections occur in 4% of kidney transplants [3–5].

To diagnose allograft rejection, tissue biopsies are considered as the gold-standard method for detecting acute and chronic immune injury, as well as other pathologies associated that may eventually lead to allograft loss. However, biopsies are invasive, costly, in rare cases they can lead to organ loss, while the readout can potentially be erroneous if a non-affected part of the kidney is sampled by chance. Therefore, proceeding with biopsies in patients of low immunological risk is sometimes criticized [6]. There is hence a strong need for non-invasive assays to detect injury in transplanted kidneys. Several studies to develop suitable biomarkers for allograft rejection have been conducted. These studies include the quantification of specific messenger RNAs in urine [7], large-scale transcriptomics analyses of peripheral blood [8], proteomics analyses of biopsies [9] and urine [10,11], and metabolomics [12] and RNA sequencing [13] of urine pellet or supernatant. Nevertheless, all non-invasive methods developed to date still have important caveats and require further improvement.

The presence of donor-specific DNA in blood was first reported in women who had a kidney and a liver transplant [14]. The measurement of cell-free donor-specific DNA in blood for a differential diagnosis of kidney injury has been suggested recently [15–17]. These studies focused on females who received a kidney from male donors by identifying the presence of DNA coding for the testis specific protein *Y-linked 1* (TSPY1) or the sex-determining region of the Y chromosome using quantitative polymerase chain reaction. With the improvement of next generation sequencing technologies, whole genome sequencing (WGS) [18,19] and targeted sequencing [20] were used for measuring donor-specific DNA for solid organ transplant rejection. However, these studies focused on heart transplants and measured cell-free donor-specific DNA in blood plasma. More importantly, these methods require the sequencing of both donor and receptor DNA which is more costly.

An algorithm for measuring donor-specific DNA in plasma of organ transplants without requiring donor or recipient genotyping was implemented by Gordon et al [21]. But this algorithm made the assumption that donor fraction is < 14%. More recently, Grskovic *et al*. used sequencing of 266 single nucleotide variants (SNVs) that discriminate best between two unrelated individuals to count reference and alternative allele frequency for estimating the donor-derived cell-free DNA fraction [22]. This method showed a high correlation between cell-free donor specific DNA levels in recipient blood and active rejection of the kidney allografts [23]. However, this method does not account for potential sequencing errors and requires *a priori* knowledge of the familial relationship between donor and recipient. Finally, a statistical method combining SNV array genotyping of donor and recipient before transplantation with recipient DNA sequencing was used to estimate recipient-derived DNA fraction in heart and lung transplants [24]. Nonetheless, this method requires SNV genotyping of donor and recipient DNA before transplantation. Most importantly, all of the previous study focused on DNA extract from blood.

The presence of donor-specific DNA in urine of kidney allograft recipients has been reported [25]. We have recently conducted a study based on RNA sequencing of tissue biopsies from kidney allograft transplants and found a correlation between the ratio of heterozygous to homozygous SNVs with the rejection phenotype [26]. Moreover, we have shown in another study that DNA methylation could be used to accurately estimate the tissue type composition in recipient urine samples. We found that the largest tissue types present in recipient urine were kidney cells and neutrophils and that donor-specific DNA fraction correlates with the kidney derived cell fraction [27]. However, we restricted the analysis on kidney recipients with urinary tract infection and BK-virus nephropathy only. Most recently, we have identified different gene signatures and pathways associated with two different type of kidney rejection using RNA-seq on transplant urine: acute T cell–mediated rejection and antibody-mediated rejection [13]. Deconvolution analysis showed a higher enrichment of immune cells in rejection stage comparing to no-rejection.

Based on the idea that the fraction of donor-specific DNA can be determined using DNA sequencing, we here hypothesize that the recipient-specific DNA fraction in urine correlates with the level of active rejection in the kidney allograft, assuming that recipient-specific DNA originates mostly from tissue-invading immune cells while donor-specific DNA stems from the allograft [28]. Inspired by methods to estimate DNA contamination in sequencing projects [29,30], we present a cost-effective method to determine the fraction of donor-specific DNA (denoted α hereafter) in urine by sequencing targeted regions. We estimate the dependence of the precision of this measure on sequencing depth and length of the targeted region. Most importantly, no prior knowledge of donor and recipient relation is required. To the best of our knowledge, this is the first method for estimating donor-specific DNA fraction in DNA mixture extracted from recipient urine. Our method provides an easy way to determine the donor-specific DNA fraction regardless of donor and recipient gender. We evaluate its applicability for the detection of kidney transplant rejection. Future applications could be routine tests of urine samples as a reference to adjust and optimize the dosage of immune suppressants in kidney transplant patients.

## Results

### In silico simulation of donor-recipient DNA mixtures

To determine the optimal sequencing parameters, we use numerical simulations. The simulation process is based on generating two different SNV-sets, merge the two sets with a predefined proportion of each set; α from set 1 and (1-α) from set 2, and then apply a likelihood function (Methods) to estimate this proportion (*observed α*). Two major parameters affect the estimation of the *observed α*: the number of sequenced SNVs (*N*) and the depth of sequencing coverage (*M*). For a range of parameters *N=*{10, 50, 100, 500, 1,000} and *M=*{10, 50, 100, 500, 1,000, 5,000, 10,000} and varying α from 0 to 0.5 in steps of 0.01, we repeated the simulation process for each *N* x *M* x α combination 1,000 times to obtain an empirical distribution of *observed α* (Fig S1).

We computed the maximum error (ε) for each combination *N* x *M* over all tested α. ε ranges between 0 (best case where *observed α* = tested α) and 0.5 (worst case where tested α = 0.5 and *observed α* = 0 or tested α = 0 and *observed α* = 0.5) (Fig 1). As expected, our simulations show that increasing both *N* and *M* improves the *observed α* estimation accuracy. Moreover, the estimation of the *observed α* is unstable when using a small number of SNVs (*N* < 100) or low coverage (*M* < 500). The prediction accuracy stabilizes above *N* > 500 and *M* >1,000.

**Fig 1.**
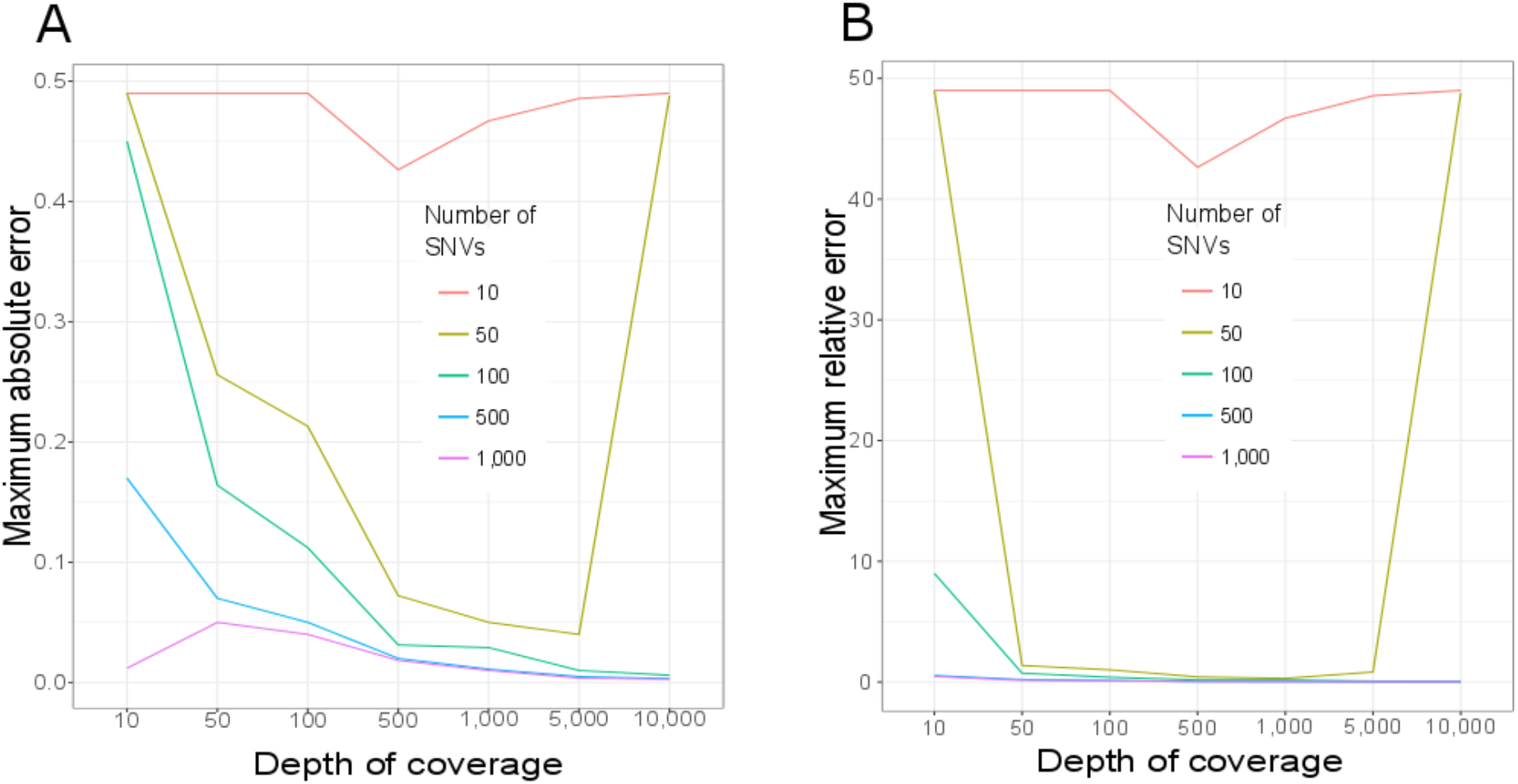
Maximum error for detecting the DNA fraction *α* in a simulated DNA sequencing experiment varying sequencing depth and number of SNVs. Maximum absolute (A) and relative (B) errors are represented. A total of 35 scenarios combining five different numbers of SNVs *N*= {10, 50, 100, 500, 1,000} and seven depth of coverage *M*= {10, 50, 100, 500, 1,000, 5,000, 10,000} were simulated (Fig S1). Represented here are maximum error observed in 1,000 simulations for every tested α ranging from 0 to 0.5 in steps of 0.01.

### Experimental estimation of α using a controlled mixture of urine from two individuals

To assess the accuracy of detecting *observed α* in a mixture of two real human urine samples, we performed a targeted sequencing of urine DNA from two healthy individuals originated from different populations; S1 a 35 years old healthy European woman and S2 a 34 years old healthy Arab woman. A total of 1,850 exonic regions from a panel targeting 93 genes known to be associated with risk of breast cancer were sequenced. These sequenced genomic regions cover 370,942 base pairs across 22 chromosomes (Table S1). A total of 51,893 bi-allelic SNVs falling in these genomic regions were present in the Exome Aggregation Consortium (ExAC) [31]. As the method works on bi-allelic SNV with different genotypes between donor and recipient, we computed for each SNV the probability of having different genotypes for two individuals (Table S1). Only 437 SNVs have a probability of having different genotypes for two individuals higher than 10%.

As a measure of quality control, we first checked the balance of reference and alternative alleles in heterozygous calls. The alternative allele frequency is expected to be around 50% in heterozygous genotypes. However, we observed the presence of SNVs with skewed alternative allele frequencies (Fig S2). We noticed the recurrence of such unbalance in every replicate of both samples (Fig S3 for examples). We investigated whether the amplification-based strategies for DNA target enrichment affect the allele dropout causing the skewed alternative allele distribution. We found that the SNVs with a skewed distribution all fall into the primer sequence regions. We therefore filtered out SNVs falling into these regions and kept the 1,000 most common SNVs in the general population. These 1,000 SNVs will be used as a SNVs panel for detecting DNA fraction in a combination of two DNA sources (*observed α*) in the rest of the study. The alternative allele frequency was balanced in these 1,000 SNVs (Fig S4). Moreover, the maximum error of estimating the *observed α* based on these 1,000 SNVs in all replicates was < 0.0034 (mean error = 0.0028 ± 0.00037).

We then mixed 90 % DNA from S1 and 10% DNA from S2 in three replicates. For each replicate, targeted DNA sequencing was performed and the *observed α* was estimated. The preparation of the mixture was based on total DNA content in the samples. However, the presence of bacterial DNA in urine samples can strongly skew the estimation of human DNA concentration measurement [32]. We assessed the actual DNA concentration of S1 and S2 in urine by considering the mean *observed α* over the three replicates to 0.053. This indicates that S1 DNA concentration is ∼19 times lower than S2 DNA concentration. Considering the estimated S1 and S2 DNA concentration, the maximum error of the *observed α* was < 3.5% in the three replicates (Fig 2). We extended the analysis to two levels of DNA mixture scenarios: (i) 70% DNA from S1 and 30% DNA from S2, (ii) 50% DNA from S1 and 50% DNA from S2. Each scenario was replicated three times and targeted DNA sequencing was performed for each replicate. The *observed α* was similar in the three replicates of all three scenarios (scenario i: mean *observed α* = 0.11 ± 0.036, scenario ii: mean *observed α* = 0.032 ± 0.00048). Considering the estimated S1 and S2 DNA concentration, the maximum error of the *observed α* was < 3.8% in all replicates of both scenarios (0.037 in scenario (i) and 0.018 in scenario (ii)) (Fig 2).

**Fig 2.**
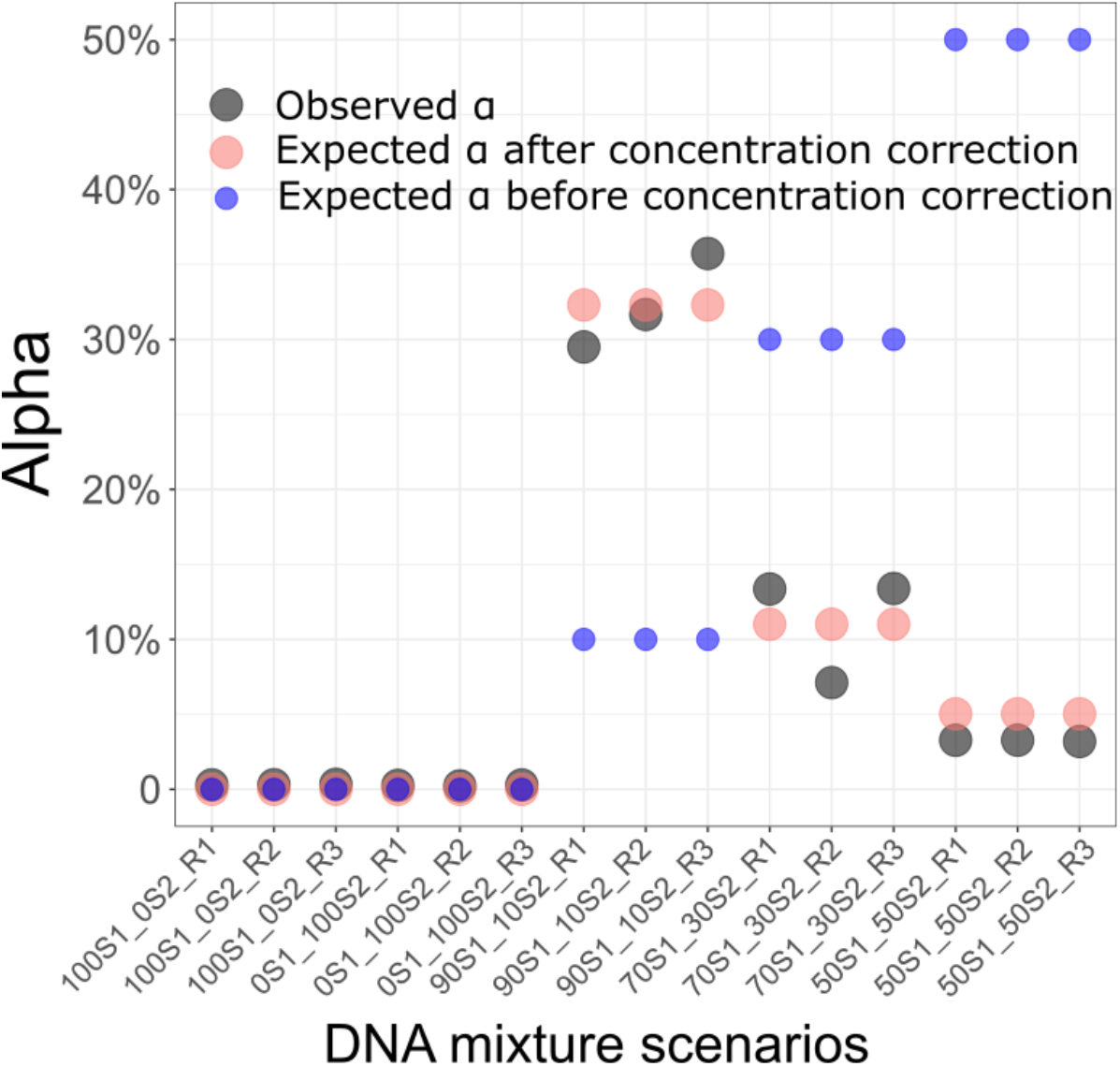
Estimation of DNA fraction (Alpha) in a combination of two healthy DNA sources. Five scenarios of DNA mixtures and three replicates for each scenario were performed. From left to right: 100% from individual S1 and 0 % from individual S2, 0% from individual S1 and 100% from individual 2, 50% from individual 1 and 50% from individual 2, 70% from individual S1 and 30% from individual S2, 90% from individual S1 and 10% from individual S2. The estimated fractions (*estimated α*) are represented by black dots. The expected fractions when DNA concentration in individual Sl was 19 times lower than DNA concentration in individual S2 are represented by red dots. The expected fractions before correction for DNA concentration are represented by blue dots.

### Simulation of the effect of family relationship and ethnicity on the estimation of α

The most challenging scenario is that of one sibling donating a kidney to another, as they share 50% of their genome. The extreme case of mono-zygotic twins, where both genomes are identical, can of course not be addressed with our method. To numerically explore this “worst case” scenario, we used whole genome sequencing data from 91 siblings [33] and then generated 100 combinations of every two siblings. Self-reported relationship was confirmed using identity by state (Fig S5). For each pair of siblings, we simulated donor and recipient DNA sequences by varying α from 0 to 0.5 in steps of 0.01 and resampling the mean coverage at 5,000 reads. The maximum absolute error was observed when the expected α = 0.07: *observed α* = 0.034 (Fig 3).

**Fig 3.**
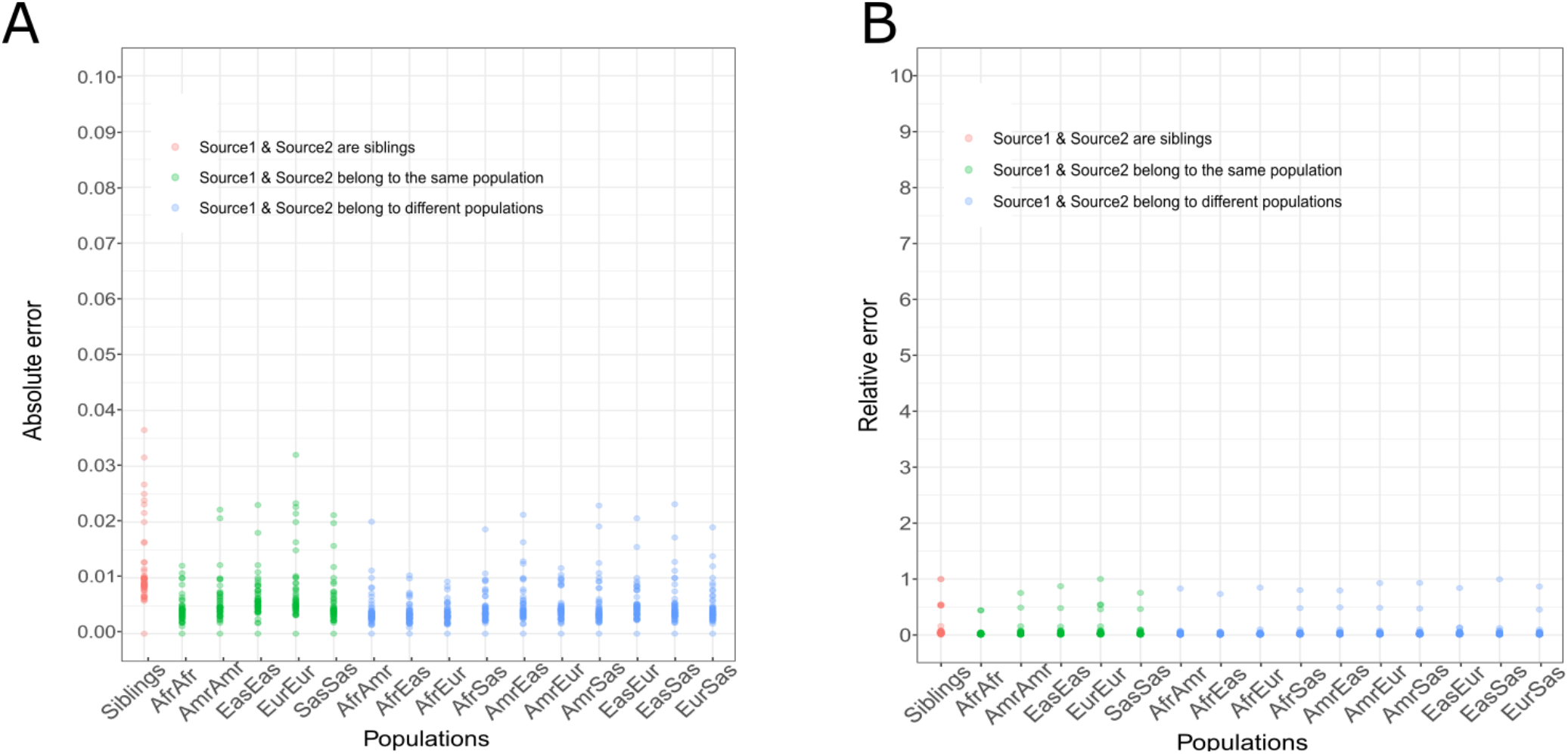
Effect on family relationship and ethnicity on detecting DNA fraction in a combination of two DNA sources. Each dot represents in A) the maximum absolute error and in B) the maximum relative error for each expected (α) from 0 to 0.5 in steps of 0.01 over 100 pairs of siblings (red), 100 pairs of individuals belonging to the same population (green) and 100 pairs of individuals belonging to different populations (blue). Afr = Africans. Amr = Americans. Eas = East Asians. Eur = Europeans. Sas = South Asians.

### Simulation of the effect of population origin on the estimation of α

As per comparison to siblings, we assessed the effect of donor and recipient ethnicity on our method. We applied our method on simulated pairs of individuals belonging to the same and to different populations of the 1,000 genomes project [34]: Africans, Americans, east Asians, Europeans and south Asians. The absolute error was < 0.04 in all scenarios (Fig 3). As expected, the absolute error was lower when the two DNA sources belong to different populations (mean maximum absolute error = 0.018 ± 0.005) than when they belong to the same population (mean maximum absolute error = 0.022 ± 0.007). Additionally, the maximum relative error was comparable in all scenarios together (mean maximum relative error = 0.836 ± 0.137) compared to when the two DNA sources belong to the same (mean maximum relative error = 0.764 ± 0.207) or different populations (mean maximum relative error = 0.856 ± 0.077). These results confirm the power of the method for detecting the DNA fraction in a combination of two DNA sources independently of the familial relationship or their ethnicity.

### Application to urine samples from clinical kidney allograft patients

To test our method in a real-case scenario, we used DNA extracted from 32 urine samples of 26 kidney allograft recipients collected at the time a diagnostic biopsy was performed and divided the sample into three groups based on their Banff classification: “Acute Tubular Injury” (ATI, N = 12), “Acute Rejection” (AR, N = 11) and “No Observed Pathology” (N = 9) (Table 1). DNA was extracted from urine and deep targeted sequencing was performed for the 32 samples. Reflecting the effect of depth of coverage on the accuracy of detecting *observed α* in simulated data, we set the mean depth of coverage to ∼ 14,000 reads. After read alignment we applied our method to estimate the donor to total DNA fraction (Fig 4).

**Table 1.** Characteristics of Kidney Transplant Recipients.

| Table 1. Characteristics of Kidney Transplant Recipients |  |  |  |  |
| --- | --- | --- | --- | --- |
| Recipient Characteristics | Patient Total<br>(N=26) | Patients<br>with AR<br>(N=8) | Patients<br>with ATI<br>(N=10) | Patients with<br>No Pathology<br>(N=8) |
| Number of Biopsy Associated<br>Urine Specimens | 32 | 12 | 11 | 9 |
| Age, years |  |  |  |  |
| Mean (SD) | 45.5 (14) | 46.1 (19) | 43.8 (14) | 47.1 (8) |
| Median | 43 | 43 | 40 | 49 |
| Min, Max | 25, 80 | 25, 80 | 29, 74 | 34, 59 |
| Gender, N (%) |  |  |  |  |
| Male | 40 (100%) | 40 (100%) | 40 (100%) | 40 (100%) |
| Race, N (%) |  |  |  |  |
| White | 8 (31%) | 3 (38%) | 3 (30%) | 2 (25%) |
| Black | 11 (42%) | 4 (50%) | 3 (30%) | 4 (50%) |
| Hispanic | 3 (11%) | 1 (12%) | 0 (0%) | 2 (25%) |
| Asian | 2 (8%) | 0 (0%) | 2 (20%) | 0 (0%) |
| Mixed | 2 (8%) | 0 (0%) | 2 (20%) | 0 (0%) |
| Cause of ESRD, N (%) |  |  |  |  |
| Diabetes | 4 (15%) | 1 (13%) | 1 (10%) | 2 (25%) |
| Hypertension | 11 (42%) | 3 (37%) | 5 (50%) | 3 (50%) |
| Glomerulonephritis | 6 (23%) | 3 (37%) | 1 (10%) | 2 (25%) |
| Polycystic Kidney Disease | 3 (12%) | 0 (0%) | 2 (20%) | 1 (11%) |
| Other | 2 (8%) | 1 (13%) | 1 (10%) | 0 (0%) |
| Prior Transplant History, N (%) | 2 (8%) | 1 (13%) | 1 (10%) | 0 (0%) |
| Donor Source, N (%) |  |  |  |  |
| Living | 17 (65%) | 4 (50%) | 7 (70%) | 6 (75%) |
| Deceased | 9 (35%) | 4 (50%) | 3 (30%) | 2 (25%) |
| Induction Therapy, N (%) |  |  |  |  |
| Antithymocyte globulin | 23 (88%) | 5 (62%) | 10 (100%) | 8 (100%) |
| IL-2 Receptor Antibody | 3 (12%) | 3 (38%) | 0 (0%) | 0 (0%) |
| Steroid Maintenance Therapy, N (%) | 10 (38%) | 4 (50%) | 2 (20%) | 4 (50%) |
| Post-Transplant Month, mean (SD) | 8.26 (12.32) | 9.79 (11.05) | 1.56 (1.64) | 15.10 (17.20) |
| Biopsy Creatinine | 3.1 (2.4) | 2.9 (1.9) | 4.4 (2.9) | 1.6 (0.25) |
| One Year Post Biopsy Creatinine | 2.2 (1.7) | 2.9 (2.9) | 2.0 (0.6) | 1.8 (0.7) |

**Fig 4.**
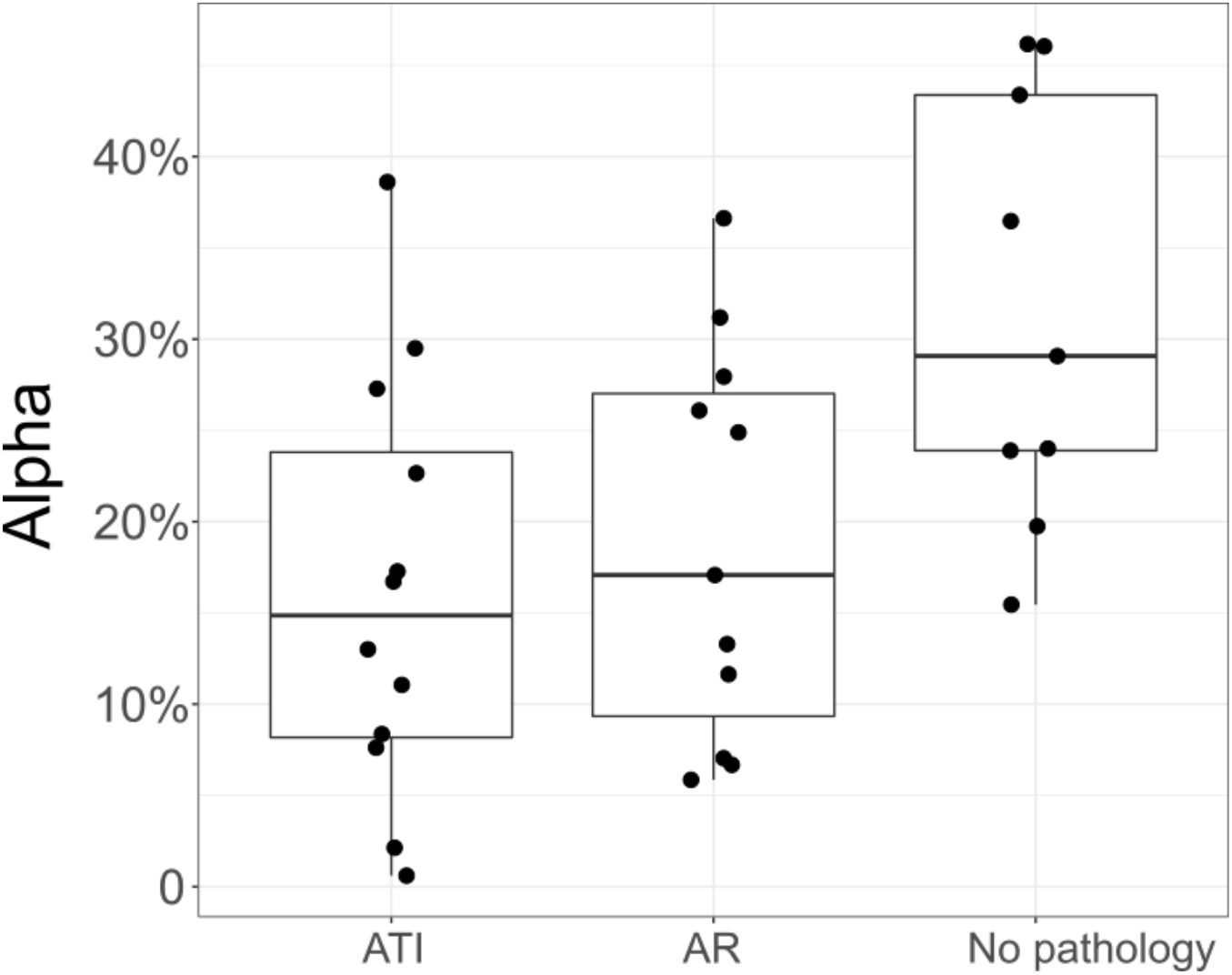
Donor to total DNA fraction in 32 real kidney allograft recipient urines. Box plots and individual data points of the estimated fraction (*observed α*) are estimated from deep urine DNA targeted sequencing. AR: Acute Rejection. ATI: Acute Tubular Injury. A statistically difference was observed between all the diagnosis phenotypes (P = 0.035, Kruskal-Wallis test). By Dunn’s test, difference in *observed α* between the two pathologies and no pathology group was statistically significant: ATI vs no-pathology: P = 0.0064 and AR vs no-pathology: P = 0.026. Pathologies pairwise comparison was not statistically significant (P > .05).

The difference of *observed α* between the diagnosis phenotypes was statistically significant (P = 0.035, Kruskal-Wallis test). We observed a significant difference when comparing the two transplant kidney pathologies ATI and AR to the No Pathology group (P = 0.0064 and P = 0.026, Dunn’s test for ATI vs no pathology and AR vs no pathology, respectively). However, no significant difference was observed in *observed α* when comparing the two pathologies ATI to AR (P = 0.31, Dunn’s test).

### Inference of donor and recipient ethnic origin

In the absence of donor and recipient genomes, it is impossible to determine whether the *observed α* represents the donor or the recipient fraction of the total DNA. However, in cases were recipient and donor gender or ethnicity differ, this issue can be addressed. The urine DNA sequencing we performed here did not target genomic regions of the Y chromosome. Thus, detecting recipient and donor gender cannot be carried out using the actual data, but could be easily amended in future sequencing panels.

To predict donor and recipient ethnicity, an estimation of both recipient and donor genotypes is needed. For each of 1,000 SNVs, we computed the fraction of the alternative to total alleles. We then used the *observed α* to compute the nine expected fractions of the alternative allele (Table 2). We then used a cost function to estimate donor and recipient genotypes that minimizes the difference between the nine expected fractions and the observed fraction of the alternative to total alleles. Based on these estimated genotypes, we applied a supervised classification method to predict the recipient and the donor ethnicity as following: as donor-specific DNA fraction has been shown to be higher in the no-pathology group [28], we supposed the *observed α* to represent the donor-to-total DNA fraction and computed the probability of donor and recipient for belonging to one of the three populations: African, East Asian and European (see Methods). Both donor and recipient are assigned to the population showing the highest probability and then compared to the self-reported ethnicity. Seven recipients and eight donors were excluded from the prediction because they belong to a mixed self-reported population or the *observed α* was ∼ 0 so the prediction of donor genotypes was impossible. The prediction was inconclusive (Probability of prediction < 70%) for 5 recipients and 8 donors. In 16 over 20 recipients (80%) and 15 over 16 donors (94%), the probability of prediction was higher than 70%. However, only one AR sample and one ATI sample have donor and recipient ethnicity mismatch for whom the prediction was conclusive. In these two samples (European donor and African recipient for both samples), the prediction was in agreement with the self-reported ethnicity. Hence, due to the small number of self-reported ethnicity mismatches, it is impossible to confirm whether the *observed α* represents the donor or the recipient DNA fraction (as *observed α* < 0.5 by definition).

**Table 2.** Expected fraction of the alternative to total alleles (reference + alternative) as a function of *observed α*.

| $g_i^R$ | $g_i^D$ | $exp_i$ |
| --- | --- | --- |
| 0 | 0 | 0 |
| 0 | 1 | $\alpha/2$ |
| 0 | 2 | $\alpha$ |
| 1 | 0 | $1 - \alpha/2$ |
| 1 | 1 | $1/2$ |
| 1 | 2 | $1 + \alpha/2$ |
| 2 | 0 | $1 - \alpha$ |
| 2 | 1 | $2 - \alpha/2$ |
| 2 | 2 | 1 |

## Discussion

Different omics technologies, including mRNA measurement by PCR [7], metabolomics [12] and RNA-sequencing [13] have been applied by our group and others to identify non-invasive biomarkers for kidney allograft rejection. Here, we present a new approach based on targeted deep-sequencing of DNA obtained from urine samples. We extended methods originally used for the assessment of DNA contamination to estimate the fraction of recipient DNA in a two-sources mixed DNA sample [29,30]. We used in silico simulations to obtain a suitable parameter range for the method to be sufficiently accurate in estimating the fraction of a two-sources DNA mixture. We then experimentally evaluated the accuracy of the estimation method using controlled mixtures of two DNA sources. Allele drop-out occurs in amplification-based target enrichment when a variant is located in a primer region and prevents primer hybridization, leading to failed amplification and allele bias [35]. Our method overcomes these unexpected artefacts due to DNA sequencing. Other algorithm for estimating the donor-specific DNA fraction requires the donor and recipient relationship information [22]. Here, we found that ethnicity and familial relationship between donor and recipient appear to have a lower impact as compared to previously presented methods

We tested our method on clinical samples from patients with and without kidney rejection events. We compared the α value obtained from urine DNA sequencing reads of kidney allograft recipients with kidney injury associated with AR and ATI. The alpha value was significantly different in patients with AR and ATI compared to those without kidney pathology. The calculation of alpha is based on the assumption that the DNA isolated form the urine is derived from the transplanted kidney where both the recipient and the donor DNA are present: Recipient DNA from the infiltrating immune cells and the donor DNA from the kidney parenchymal cells. Indeed, we have recently shown in kidney recipients with donor-recipient gender mismatch by counting Y chromosome-derived cell free DNA that donor-specific DNA fraction was lower in recipient with UTI as comparing to the no UTI and higher in recipients with BKVN comparing to the no BKVN [28]. Thus, our approach might be considered as a potential new diagnostic signature measured in urine specimens.

We were not able to assert whether the recipient urine DNA is mostly donor’s or recipient’s. Studies have shown that both AR and ATI are associated with allograft damage indicating that there will be some donor DNA in the urine. But AR is also associated with recipient immune cell infiltration while ATI is not [36]. Thus, the fraction of recipient to donor cells in the urine should be higher for AR compared to ATI and perhaps should be just the opposite with AR showing a fraction of donor to recipient of much lower than 0.5 and ATI showing a fraction of donor to recipient of much greater than 0.5. To address this, a future complementary analysis on a bigger sample having donor and recipient ethnicity and/or gender mismatches will be worthwhile.

## Methods

### Algorithm

The algorithm is inspired by the contamination estimation in DNA sequencing method [29,30]. We hypothesize that recipient urine contains a mix of recipient and donor DNA. Let *N* be the number of bi-allelic SNVs sequenced from recipient urine DNA and each SNV *i* is covered by *M*_*i*_ reads. Let g_i_^R^ and g_i_^D^ be the genotype of recipient and donor at the SNV *i*, respectively. Both g_i_^R^ and g_i_^D^ are unknown. Limiting the analysis on bi-allelic SNVs only leads to three possible genotypes for recipient and donor at each SNV *i*: g_i_^R^ (g_i_^D^)= {0, 1, 2} where 0 = homozygous wild type, 1 = heterozygous and 2 = homozygous for the alternative allele. The likelihood of the donor-specific DNA fraction (α) will be:

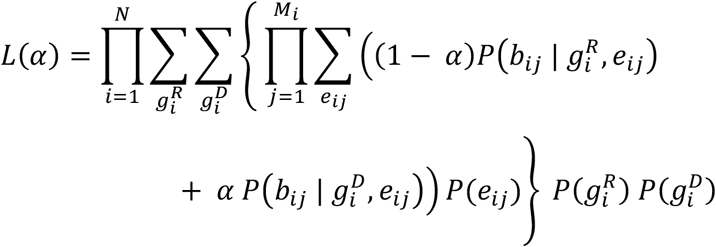

Where *b*_*ij*_ represents the read *j* covering the SNV *i* and *e*_*ij*_ represents the sequencing error of SNV *i* at the read *j* : P(*e*_*ij*_ = 1) = 10^−*Qij/10*^ and P(*e*_*ij*_ = 0) = 1-P(*e*_*ij*_ = 1) and *Q*_*ij*_ represents the minimum between the base quality of the read *j* at the position of the variant *i* and the mapping quality of the read *j*. The probability of *b*_*ij*_ conditioned to the recipient (donor) genotype g_i_^R^ (g_i_^D^) and the sequencing error *e*_*ij*_ is described in Table 3. Finally, we used the simulated annealing approach together with a grid search to find α that maximizes the likelihood function [37]. The method was implemented in a Python script.

**Table 3.** The probability of read *b*_*ij*_ carrying the reference (a), alternative (A) or a different allele (e) conditioned to the recipient (donor) genotype g_i_^R^ (g_i_^D^) and the sequencing error *e*_*ij*_.

| | $g_i^R (g_i^D) = 0$ | | $g_i^R (g_i^D) = 1$ | | $g_i^R (g_i^D) = 2$ | |
| --- | --- | --- | --- | --- | --- | --- |
| | $e_{ij} = 0$ | $e_{ij} = 1$ | $e_{ij} = 0$ | $e_{ij} = 1$ | $e_{ij} = 0$ | $e_{ij} = 1$ |
| $P(b_{ij} = a)$ | 1 | 0 | 1/2 | 1/6 | 0 | 1/3 |
| $P(b_{ij} = A)$ | 0 | 1/3 | 1/2 | 1/6 | 1 | 0 |
| $P(b_{ij} = e)$ | 0 | 2/3 | 0 | 2/3 | 0 | 2/3 |

As the likelihood function for estimating α requires a balance in alternative/reference allele distribution in heterozygous calls for both recipient g_i_^R^ and donor g_i_^D^ genotypes, very deep recipient urine DNA sequencing will provide this allele balance (Table 4).

**Table 4.** The probability and the 99% interval confidence of a perfect allele balance in a heterozygous call as a function of depth of coverage *M*. A total of 10,000 simulations were performed for each proposed *M*.

| $M$ | $P(\text{alt}/(\text{ref}+\text{alt}) = 0.5)$ | 99% confidence interval |
| --- | --- | --- |
| 10 | 0.24 | [0.10, 0.90] |
| 50 | 0.11 | [0.34, 0.66] |
| 100 | 0.08 | [0.39, 0.62] |
| 500 | 0.18 | [0.45, 0.55] |
| 1,000 | 0.27 | [0.46, 0.54] |
| 5,000 | 0.53 | [0.48, 0.52] |
| 10,000 | 0.69 | [0.49, 0.51] |

### Simulated SNVs based on general population structure

To assess the effect of number of SNVs (*N*) and mean depth of coverage (*M*) on the allele balance and thus the prediction accuracy of the likelihood function, we simulated 2 independent SNV-sets each set containing *N* common SNVs (minor allele frequency ≥ 5%) and is covered on average by *M* reads. We varied *N* and *M* in 35 scenarios where *N*={10, 50, 100, 500 and 1,000} and *M*={10, 50, 100, 500, 1,000, 5,000 and 10,000}. We merged α reads from set1 and 1-α from set2 randomly generating a combined SNV-set and applied the likelihood function on the combined SNV-set to estimate the *observed α*. Because *L(α) = L(1-α)*, we restrict 0 ≤ α ≤ 0.5 in steps of 0.01 generating 51 scenarios. A thousand replicates for each scenario and for each α were performed to obtain an empirical distribution.

### Sequencing of urine DNA from a pair of healthy individuals

We extracted DNA from urine of two healthy individuals; S1, a 30 years old European woman and S2 a 30 years old Arab woman using the Qiagen® Allprep Mini Kit. extraction kit. The DNA concentration was similar for the two individuals: 35ng/μl. We mixed DNA from S1 and S2 to achieve 5 scenarios: i) 100% from S1, ii) 100% from S2, iii) 90% from S1 and 10% from S2, iv) 70% from S1 and 30% from S2, v) 50% from S1 and 50% from S2 and each scenario was replicated three times. We performed deep targeted DNA sequencing on each replicate. GeneReadDNA Seq Targeted Pannels V2; Human Breast Cancer Panel (Qiagen, USA) was used to perform target enrichment by multiplex PCR. The breast cancer panel consists of four primer pools yielding 2,915 amplicons. Briefly, 40ng of each gDNAs was amplified using PCR reagents with 4 primer pool mixes following the manufacturer’s protocol. After the completion of the 4 PCR reactions, the 4 products were combined and the enriched DNA was purified using Agencourt AMPure XP beads (Beckman Coulter, USA). The concentration and the size of the purified amplicons were determined using Qubit 2.0 Fluerometer dsDNA BR assay kit (LifeTechnologies, USA) and Agilent BioAnalyzer 2100 High-Sensitivity DNA kit (Agilent Technologies, USA). A total amount of 80-160 ng of purified enriched DNA was used as template to generate NGS libraries. The NGS libraries were prepared using NEBNEXT Ultra II DNA Library Prep Kit (New England Biolabs, USA) and NEXTflex DNA Barcodes (Bio Scientific, USA). All library preparation steeps were performed according to the manufacturer’s protocol. The size and quality of the final libraries were analyzed using Agilent BioAnalyzer 2100 with 1000 DNA kit (Agilent Technologies, USA). The quantified libraries were then normalized, pooled, and spiked with 5% PhiX control library (Illumina, USA). Finally, thepooled libraries were sequenced on a singles lane of Illumina Hiseq 4000 (Illumina, USA) paired-end 150 bp run.

Obtained reads were aligned to the human genome reference hg19 using bwa [38]. A total of 51,893 bi-allelic SNVs from the Exac project are included in the targeted genomic regions. The method works only on SNVs with different genotype between donor and recipient. Under Hardy Weinberg assumption, we assessed the probability of having a different genotype for each SNV *i* as:

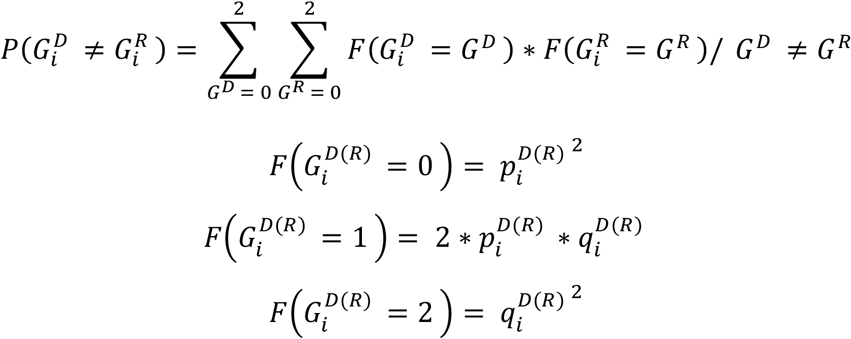

Where 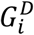 and 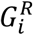 represent the donor and recipient genotype, respectively. 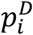 and 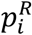 represent the donor and recipient reference allele frequency, respectively. 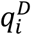 and 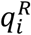 represent the donor and recipient alternative allele frequency, respectively.

To avoid allele dropout due to primer annealing region, we filtered out 24,237 SNVs falling at the primer sequencing regions [35]. From the 27,656 remaining SNVs, we selected the 1,000 most common and applied the likelihood function after filtering out the reads carrying the variant at the last 20 base pairs [39].

### Simulated SNVs in pairs of individuals from the same and different ethnicity

We used individuals from the 1,000 genomes project phase 3 representing five major populations: AFR, AMR, EAS, EUR and SAS [34]. We randomly selected two individuals and aimed to cover all possible situations; five cases where Individual1 and individual2 belong to the same population and ten cases where individual1 and individual2 belong to different populations. We extracted the 1,000 SNVs described previously from Individuals1 and Individual2 and then merged α reads from individual1 and 1-α from individual2 generating a combined SNV-set. We varied α from 0 to 0.5 in steps of 0.01 and fixed the mean depth of coverage at *M* = 5,000. We applied the likelihood function to assess α for each combined SNV-set generated. We repeated the individual selecting process 100 times to obtain an empirical distribution for each situation.

### Simulated SNVs in pairs of real siblings

We performed WGS on Illumina HiSeq 2500 sequencer of 91 Qatari siblings from 27 nuclear families containing at least 2 siblings at up to 7 siblings [33]. Reads were aligned to the hg19 reference genome using bwa [38]. Sequence alignment files were filtered and genotypes were called using the Genome Analysis Tool Kit best practices pipeline and variants were called using HaplotypeCaller [40,41]. We used Plink identity by state to confirm the familial relationship [42] (Fig S5). We extracted the 1,000 SNVs described previously from each sibling and merged each couple generating 100 combined SNV-sets. We applied the likelihood function to assess α for each combined SNV-set generated by varying α from 0 to 0.5 with a step of 0.01 while the mean depth of coverage was set at *M* = 5,000.

### Donor-specific DNA fraction in real kidney recipient urine DNA

We studied 32 biopsy matched urine specimens collected from 26 kidney allograft recipients who were enrolled in the IRB approved study protocol entitled “Use of urine PCR to evaluate renal allograft status” at Weill Cornell Medicine-New York Presbyterian Hospital. Kidney allograft biopsies were classified as acute rejection (n= 12), acute tubular necrosis (n=11) and normal histology (n=9) using the Banff 2017 schema [43] (Table 1). DNA was extracted from urinary cells and deep targeted DNA sequencing was performed on all samples. Briefly, 50cc of fresh urine was centrifuged at 2,000g for 30 minutes at room temperature and the urine cell pellet was harvested after removing the supernatant. After washing the urine cell pellet with 1ml PBS, the cells were lysed using 350ul of Buffer RLT from Qiagen® and DNA was isolated from the cell pellet using Allprep DNA/RNA/Protein Mini Kit from Qiagen®. Total DNA was quantified using the NanoDrop™ Spectrophotometer. DNA sequencing was performed as previously described for the pair of healthy individuals. Obtained reads were aligned to the human genome reference hg19 using bwa [38]. We filtered out low quality reads using an in-house python script. We applied the likelihood function on the 1,000 SNV-set to estimate the recipient-specific DNA fraction. Nonparametric Kruskal-Wallis test was applied to assess the correlation between *observed α* and all the diagnosis phenotypes. Dunn’s function was applied to test the pairwise association. R software was used for statistical tests and generating graphs [44].

### Ethnicity estimation for donor and recipient

We combined the *observed α* in the kidney transplant patients with a cost function to predict the genotype of both recipient (g_i_^R^) and donor (g_i_^D^) at each SNV *i*. First, we computed the expected fraction of the alternative to the total allele (reference + alternative) *exp*_*i*_ to all 9 possible combinations of g_i_^R^ and g_i_^D^ (Table 2). The observed fraction of the alternative to total alleles (*obs*_*i*_) at the SNV *i* is defined by:

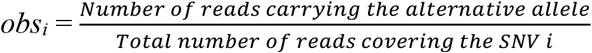

Then, we used the cost function to determine g_i_^R^ and g_i_^D^ that minimizes the difference between the 9 expected (*exp*_*i*_) and the observed (*obs*_*i*_) fraction of the alternative to total alleles:

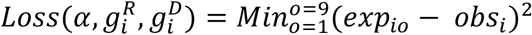

Once g_i_^R^ and g_i_^D^ estimated, we performed a partial least square analysis (PLS) using 3 subpopulations from the 1,000 genomes project African, east Asian and European populations using the mixOmics R package [45] and then predicted the ethnicity of donor and recipient in the real kidney transplant samples. The leave 2 out 1,000 fold cross validation showed the highest prediction accuracy = 81.6% reached when using the Yoruba in Ibadan in Nigeria, the Southern Han Chinese and the Toscani in Italia amongst the African, east Asian and European subpopulations (Fig S6). We excluded the American and the south Asian populations because using 1,000 SNVs only is too small to perform a reliable PLS on 5 populations where the highest cross validation prediction accuracy was too low on five populations: 54.8% (Fig S6). Additionally, none of the donors and recipients involved in the study belongs to the south Asian or the American populations.

## Data Availability

Data cannot be shared publicly because of ethical restrictions. Data are available from the Weill Cornell Medicine Institutional Data Access / Ethics Committee (contact via) for researchers who meet the criteria for access to confidential data.

## Acknowledgments

This work was supported by the Biomedical Research Program at Weill Cornell Medicine in Qatar, a program funded by the Qatar Foundation. This work was fund, in part, by awards from the NIH to MS (NIH MERIT Award, R37AI051652), TM (K08DK087824), and Weill Cornell Medical College (Clinical and Translational Science Center Award UL1TR000457). Lastly, this work was supported, in part, by the Qatar National research Fund (National Priorities Research Program grant # NPRP12S-0227-190173).

## Ethics Statement

Kidney transplant recipients reported herein provided written informed consent to participate in Weill Cornell Medicine IRB approved study protocol entitled “Use of urine PCR to evaluate renal allograft status” and the informed consent was obtained before collection of urine specimen and inclusion in the study protocol. The participants provided consent for storage of biospecimens and use of these specimens in future research. The clinical and research activities that we report here are consistent with the principles of the “Declaration of Istanbul on Organ Trafficking and Transplant Tourism” and the “World Medical Association Declaration of Helsinki on Ethical Principles for Medical Research Involving Human Subjects” [46,47].

## References

1. Hume DM, Merrill JP, Miller BF, Thorn GW. Experiences with renal homotransplantation in the human: report of nine cases. J Clin Invest. 1955;34: 327–382. doi:10.1172/JCI103085

2. Tsai M-K, Wu F-LL, Lai I-R, Lee C-Y, Hu R-H, Lee P-H. Decreased acute rejection and improved renal allograft survival using sirolimus and low-dose calcineurin inhibitors without induction therapy. Int J Artif Organs. 2009;32: 371–380.

3. Nankivell BJ, Kuypers DRJ. Diagnosis and prevention of chronic kidney allograft loss. Lancet Lond Engl. 2011;378: 1428–1437. doi:10.1016/S0140-6736(11)60699-5

4. Jalalzadeh M, Mousavinasab N, Peyrovi S, Ghadiani MH. The impact of acute rejection in kidney transplantation on long-term allograft and patient outcome. Nephro-Urol Mon. 2015;7: e24439. doi:10.5812/numonthly.24439

5. McDonald S, Russ G, Campbell S, Chadban S. Kidney transplant rejection in Australia and New Zealand: relationships between rejection and graft outcome. Am J Transplant Off J Am Soc Transplant Am Soc Transpl Surg. 2007;7: 1201–1208. doi:10.1111/j.1600-6143.2007.01759.x

6. Rush D. Can protocol biopsy better inform our choices in renal transplantation? Transplant Proc. 2009;41: S6–8. doi:10.1016/j.transproceed.2009.06.092

7. Suthanthiran M, Schwartz JE, Ding R, Abecassis M, Dadhania D, Samstein B, et al. Urinary-cell mRNA profile and acute cellular rejection in kidney allografts. N Engl J Med. 2013;369: 20–31. doi:10.1056/NEJMoa1215555

8. Li L, Khatri P, Sigdel TK, Tran T, Ying L, Vitalone MJ, et al. A peripheral blood diagnostic test for acute rejection in renal transplantation. Am J Transplant Off J Am Soc Transplant Am Soc Transpl Surg. 2012;12: 2710–2718. doi:10.1111/j.1600-6143.2012.04253.x

9. Nakorchevsky A, Hewel JA, Kurian SM, Mondala TS, Campbell D, Head SR, et al. Molecular mechanisms of chronic kidney transplant rejection via large-scale proteogenomic analysis of tissue biopsies. J Am Soc Nephrol JASN. 2010;21: 362–373. doi:10.1681/ASN.2009060628

10. Sigdel TK, Kaushal A, Gritsenko M, Norbeck AD, Qian W-J, Xiao W, et al. Shotgun proteomics identifies proteins specific for acute renal transplant rejection. Proteomics Clin Appl. 2010;4: 32–47. doi:10.1002/prca.200900124

11. Ling XB, Sigdel TK, Lau K, Ying L, Lau I, Schilling J, et al. Integrative urinary peptidomics in renal transplantation identifies biomarkers for acute rejection. J Am Soc Nephrol JASN. 2010;21: 646–653. doi:10.1681/ASN.2009080876

12. Suhre K, Schwartz JE, Sharma VK, Chen Q, Lee JR, Muthukumar T, et al. Urine Metabolite Profiles Predictive of Human Kidney Allograft Status. J Am Soc Nephrol JASN. 2016;27: 626–636. doi:10.1681/ASN.2015010107

13. Verma A, Muthukumar T, Yang H, Lubetzky M, Cassidy MF, Lee JR, et al. Urinary cell transcriptomics and acute rejection in human kidney allografts. JCI Insight. 2020;5. doi:10.1172/jci.insight.131552

14. Lo YM, Tein MS, Pang CC, Yeung CK, Tong KL, Hjelm NM. Presence of donor-specific DNA in plasma of kidney and liver-transplant recipients. Lancet Lond Engl. 1998;351: 1329–1330.

15. García Moreira V, Prieto García B, Baltar Martín JM, Ortega Suárez F, Alvarez FV. Cell-free DNA as a noninvasive acute rejection marker in renal transplantation. Clin Chem. 2009;55: 1958–1966. doi:10.1373/clinchem.2009.129072

16. Sigdel TK, Vitalone MJ, Tran TQ, Dai H, Hsieh S-C, Salvatierra O, et al. A rapid noninvasive assay for the detection of renal transplant injury. Transplantation. 2013;96: 97–101. doi:10.1097/TP.0b013e318295ee5a

17. Macher HC, Suárez-Artacho G, Guerrero JM, Gómez-Bravo MA, Álvarez-Gómez S, Bernal-Bellido C, et al. Monitoring of transplanted liver health by quantification of organ-specific genomic marker in circulating DNA from receptor. PloS One. 2014;9: e113987. doi:10.1371/journal.pone.0113987

18. Snyder TM, Khush KK, Valantine HA, Quake SR. Universal noninvasive detection of solid organ transplant rejection. Proc Natl Acad Sci U S A. 2011;108: 6229–6234. doi:10.1073/pnas.1013924108

19. De Vlaminck I, Valantine HA, Snyder TM, Strehl C, Cohen G, Luikart H, et al. Circulating cell-free DNA enables noninvasive diagnosis of heart transplant rejection. Sci Transl Med. 2014;6: 241ra77. doi:10.1126/scitranslmed.3007803

20. Hidestrand M, Tomita-Mitchell A, Hidestrand PM, Oliphant A, Goetsch M, Stamm K, et al. Highly sensitive noninvasive cardiac transplant rejection monitoring using targeted quantification of donor-specific cell-free deoxyribonucleic acid. J Am Coll Cardiol. 2014;63: 1224–1226. doi:10.1016/j.jacc.2013.09.029

21. Gordon PMK, Khan A, Sajid U, Chang N, Suresh V, Dimnik L, et al. An Algorithm Measuring Donor Cell-Free DNA in Plasma of Cellular and Solid Organ Transplant Recipients That Does Not Require Donor or Recipient Genotyping. Front Cardiovasc Med. 2016;3: 33. doi:10.3389/fcvm.2016.00033

22. Grskovic M, Hiller DJ, Eubank LA, Sninsky JJ, Christopherson C, Collins JP, et al. Validation of a Clinical-Grade Assay to Measure Donor-Derived Cell-Free DNA in Solid Organ Transplant Recipients. J Mol Diagn JMD. 2016;18: 890–902. doi:10.1016/j.jmoldx.2016.07.003

23. Bloom RD, Bromberg JS, Poggio ED, Bunnapradist S, Langone AJ, Sood P, et al. Cell-Free DNA and Active Rejection in Kidney Allografts. J Am Soc Nephrol JASN. 2017. doi:10.1681/ASN.2016091034

24. Sharon E, Shi H, Kharbanda S, Koh W, Martin LR, Khush KK, et al. Quantification of transplant-derived circulating cell-free DNA in absence of a donor genotype. PLoS Comput Biol. 2017;13: e1005629. doi:10.1371/journal.pcbi.1005629

25. Zhong XY, Hahn D, Troeger C, Klemm A, Stein G, Thomson P, et al. Cell-free DNA in urine: a marker for kidney graft rejection, but not for prenatal diagnosis? Ann N Y Acad Sci. 2001;945: 250–257.

26. Thareja G, Yang H, Hayat S, Mueller FB, Lee JR, Lubetzky M, et al. Single nucleotide variant counts computed from RNA sequencing and cellular traffic into human kidney allografts. Am J Transplant Off J Am Soc Transplant Am Soc Transpl Surg. 2018;18: 2429–2442. doi:10.1111/ajt.14870

27. Cheng AP, Burnham P, Lee JR, Cheng MP, Suthanthiran M, Dadhania D, et al. A cell-free DNA metagenomic sequencing assay that integrates the host injury response to infection. Proc Natl Acad Sci U S A. 2019;116: 18738–18744. doi:10.1073/pnas.1906320116

28. Burnham P, Dadhania D, Heyang M, Chen F, Westblade LF, Suthanthiran M, et al. Urinary cell-free DNA is a versatile analyte for monitoring infections of the urinary tract. Nat Commun. 2018;9: 2412. doi:10.1038/s41467-018-04745-0

29. Jun G, Flickinger M, Hetrick KN, Romm JM, Doheny KF, Abecasis GR, et al. Detecting and estimating contamination of human DNA samples in sequencing and array-based genotype data. Am J Hum Genet. 2012;91: 839–848. doi:10.1016/j.ajhg.2012.09.004

30. Flickinger M, Jun G, Abecasis GR, Boehnke M, Kang HM. Correcting for Sample Contamination in Genotype Calling of DNA Sequence Data. Am J Hum Genet. 2015;97: 284–290. doi:10.1016/j.ajhg.2015.07.002

31. Fu W, O’Connor TD, Jun G, Kang HM, Abecasis G, Leal SM, et al. Analysis of 6,515 exomes reveals the recent origin of most human protein-coding variants. Nature. 2013;493: 216–220. doi:10.1038/nature11690

32. Li X, Wu Y, Zhang L, Cao Y, Li Y, Li J, et al. Comparison of three common DNA concentration measurement methods. Anal Biochem. 2014;451: 18–24. doi:10.1016/j.ab.2014.01.016

33. Kumar P, Al-Shafai M, Al Muftah WA, Chalhoub N, Elsaid MF, Aleem AA, et al. Evaluation of SNP calling using single and multiple-sample calling algorithms by validation against array base genotyping and Mendelian inheritance. BMC Res Notes. 2014;7: 747. doi:10.1186/1756-0500-7-747

34. 1000 Genomes Project Consortium, Auton A, Brooks LD, Durbin RM, Garrison EP, Kang HM, et al. A global reference for human genetic variation. Nature. 2015;526: 68–74. doi:10.1038/nature15393

35. Gray PN, Dunlop CLM, Elliott AM. Not All Next Generation Sequencing Diagnostics are Created Equal: Understanding the Nuances of Solid Tumor Assay Design for Somatic Mutation Detection. Cancers. 2015;7: 1313–1332. doi:10.3390/cancers7030837

36. S O, Jf B, Pa K, Ac W, Lc R, K S. Primary acute renal failure (“acute tubular necrosis”) in the transplanted kidney: morphology and pathogenesis. In: Medicine [Internet]. Medicine (Baltimore); May 1989 [cited 21 Sep 2020]. doi:10.1097/00005792-198905000-00005

37. Kirkpatrick S, Gelatt CD, Vecchi MP. Optimization by simulated annealing. Science. 1983;220: 671–680. doi:10.1126/science.220.4598.671

38. Li H, Durbin R. Fast and accurate long-read alignment with Burrows-Wheeler transform. Bioinforma Oxf Engl. 2010;26: 589–595. doi:10.1093/bioinformatics/btp698

39. CASPER: context-aware scheme for paired-end reads from high-throughput amplicon sequencing. - PubMed - NCBI. [cited 31 May 2018]. Available: https://www.ncbi.nlm.nih.gov/pubmed/25252785

40. McKenna A, Hanna M, Banks E, Sivachenko A, Cibulskis K, Kernytsky A, et al. The Genome Analysis Toolkit: a MapReduce framework for analyzing next-generation DNA sequencing data. Genome Res. 2010;20: 1297–1303. doi:10.1101/gr.107524.110

41. Belkadi A, Bolze A, Itan Y, Cobat A, Vincent QB, Antipenko A, et al. Whole-genome sequencing is more powerful than whole-exome sequencing for detecting exome variants. Proc Natl Acad Sci U S A. 2015;112: 5473–5478. doi:10.1073/pnas.1418631112

42. Purcell S, Neale B, Todd-Brown K, Thomas L, Ferreira MAR, Bender D, et al. PLINK: a tool set for whole-genome association and population-based linkage analyses. Am J Hum Genet. 2007;81: 559–575. doi:10.1086/519795

43. C R, N S, M CG, M H, Kj H, C H, et al. A 2018 Reference Guide to the Banff Classification of Renal Allograft Pathology. In: Transplantation [Internet]. Transplantation; Nov 2018 [cited 21 Sep 2020]. doi:10.1097/TP.0000000000002366

44. R: a language and environment for statistical computing. [cited 8 Nov 2018]. Available: https://www.gbif.org/tool/81287/r-a-language-and-environment-for-statistical-computing

45. Cao K-AL, Rohart F, Gonzalez I, Gautier SD with key contributors B, Monget FB and contributions from P, Coquery J, et al. mixOmics: Omics Data Integration Project. 2018. Available: https://CRAN.R-project.org/package=mixOmics

46. Participants in the International Summit on Transplant Tourism and Organ Trafficking Convened by The Transplantation Society and International Society of Nephrology in Istanbul T. The Declaration of Istanbul on Organ Trafficking and Transplant Tourism. Clin J Am Soc Nephrol CJASN. 2008;3: 1227. doi:10.2215/CJN.03320708

47. World Medical Association Declaration of Helsinki: ethical principles for medical research involving human subjects. In: JAMA [Internet]. JAMA; 27 Nov 2013 [cited 21 Sep 2020]. doi:10.1001/jama.2013.281053

48. Robinson JT, Thorvaldsdóttir H, Winckler W, Guttman M, Lander ES, Getz G, et al. Integrative genomics viewer. Nat Biotechnol. 2011;29: 24–26. doi:10.1038/nbt.1754

