## Supplemental Figures and Table for "Deep sequencing of DNA from urine of kidney allograft recipients to estimate the donor-specific DNA fraction": Supplemental data.pdf

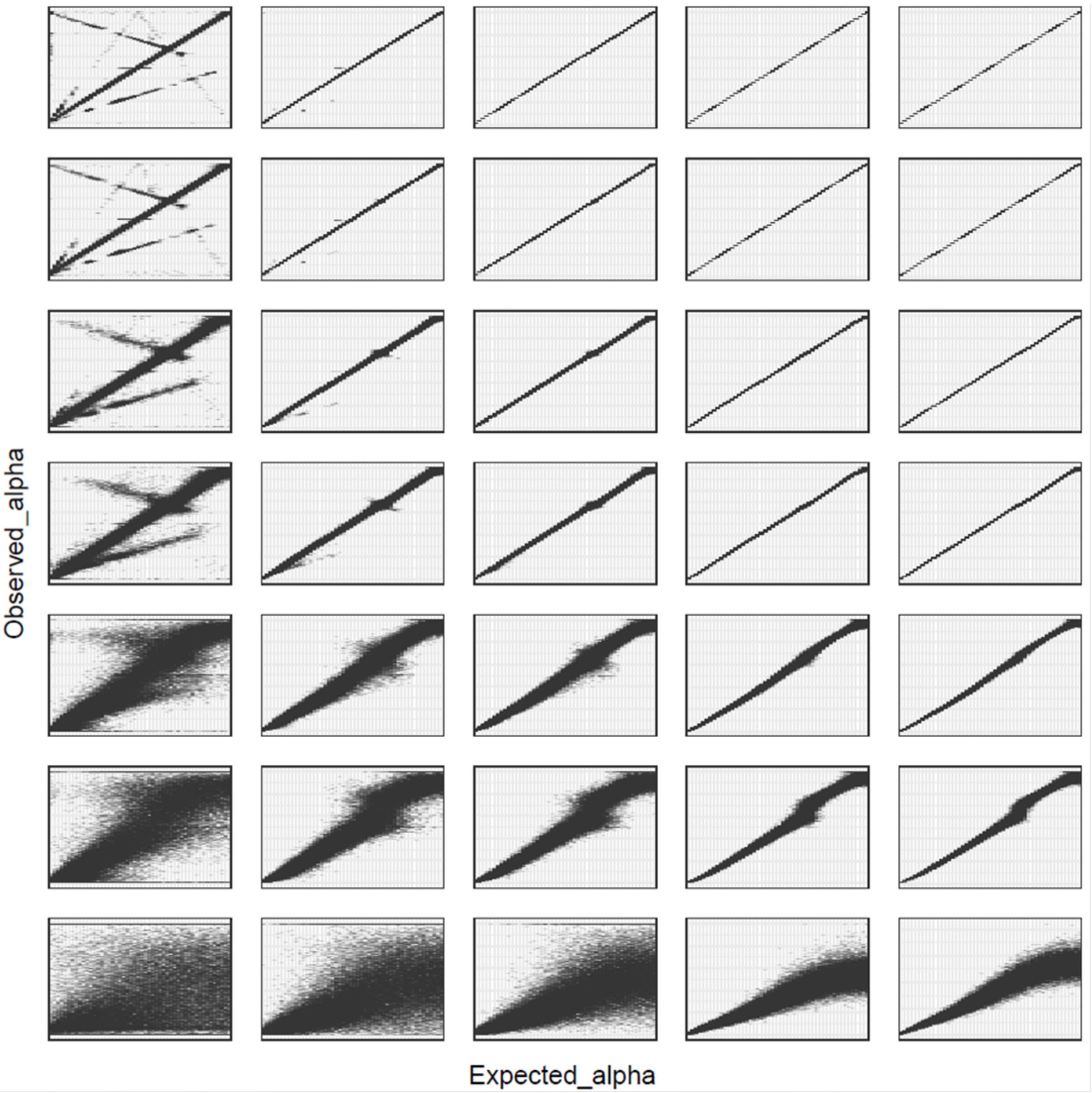


**Fig S1. Distribution of DNA fraction estimation in a combination of 2 simulated DNA sources.** A total of 35 scenarios are represented here; from left to right, number of SNVs simulated: 10, 50, 100, 500, 1,000; from down to top, mean depth of coverage: 10, 50, 100, 500, 1,000, 5,000 and 10,000. For each scenario, 51 fractions were tested from 0 to 0.5 in steps of 0.01 and the observed α (Y axis) for 1,000 simulations is shown for every α tested (X axis).

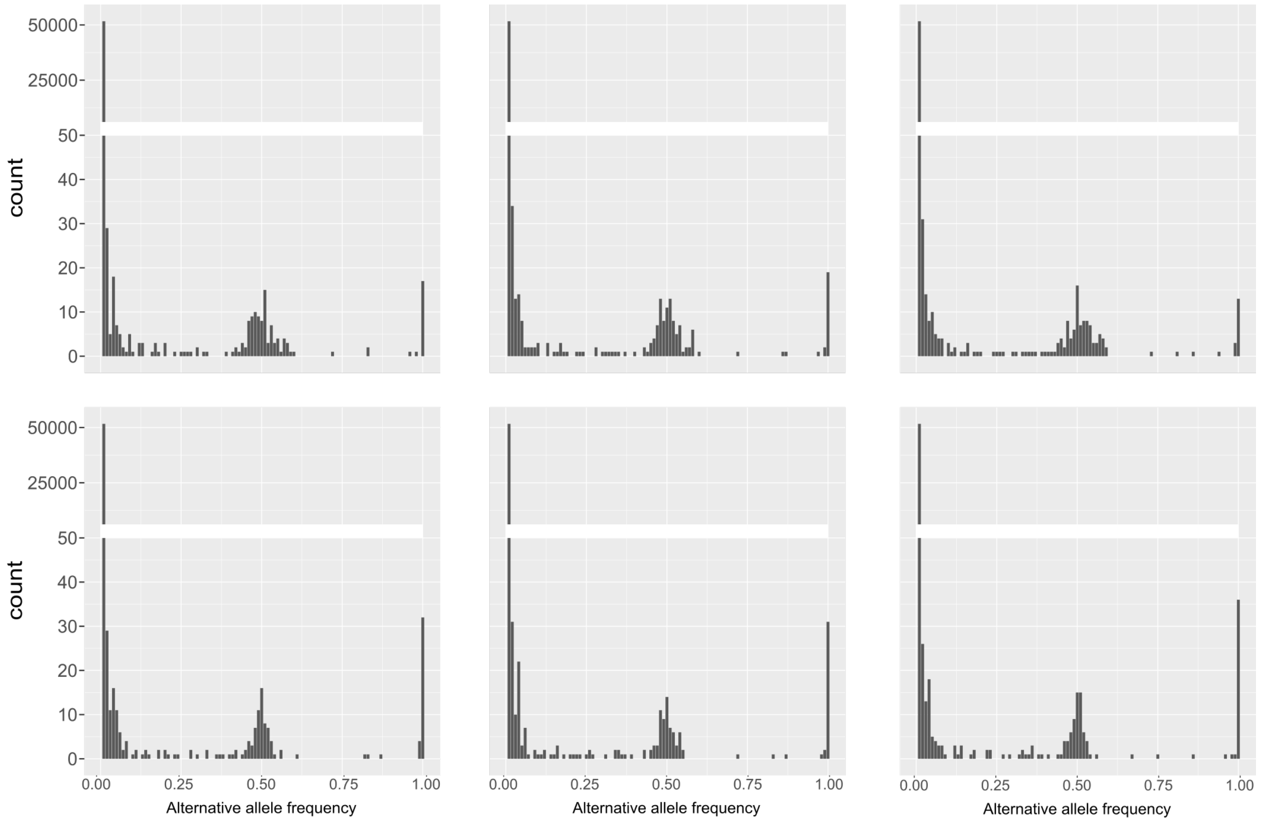


**Fig S2. Distribution of the alternative allele frequency in targeted sequencing of urine DNA from two healthy individuals.** Three replicates for individual 1 (top) and for individual 2 (Bottom) were sequenced. A total of 51,893 SNVs identified in the ExAC project are presented here. Expected alternative allele frequency ~ 0, 0.5 and 1 for homozygous wild type, heterozygous and homozygous for the alternative allele, respectively. For both individuals, the presence of SNVs with unbalanced alternative allele frequency is observed.

16

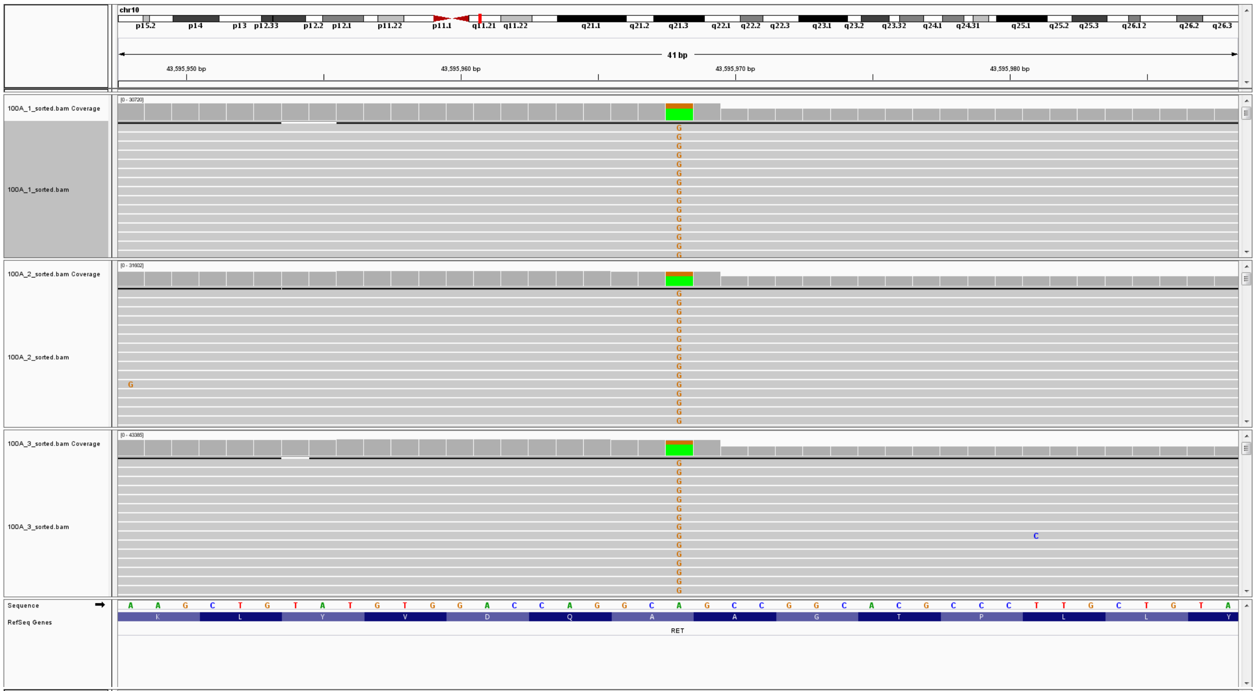
chr10
41 bp
43,595,950 bp
43,595,960 bp
43,595,970 bp
43,595,980 bp
100A_1_sorted.bam Coverage
100A_1_sorted.bam
100A_2_sorted.bam Coverage
100A_2_sorted.bam
100A_3_sorted.bam Coverage
100A_3_sorted.bam
Sequence
RefSeq Genes
RET

17

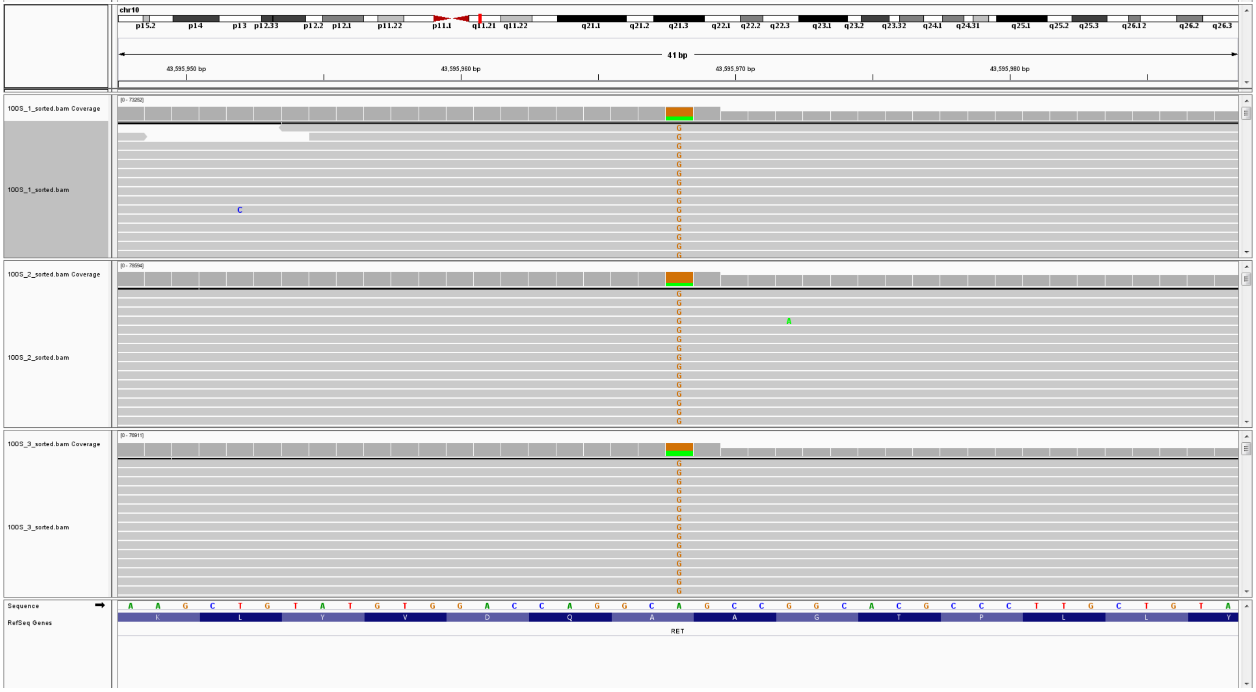
chr10
41 bp
43,595,950 bp
43,595,960 bp
43,595,970 bp
43,595,980 bp
100S_1_sorted.bam Coverage
100S_1_sorted.bam
100S_2_sorted.bam Coverage
100S_2_sorted.bam
100S_3_sorted.bam Coverage
100S_3_sorted.bam
Sequence
RefSeq Genes
RET

18

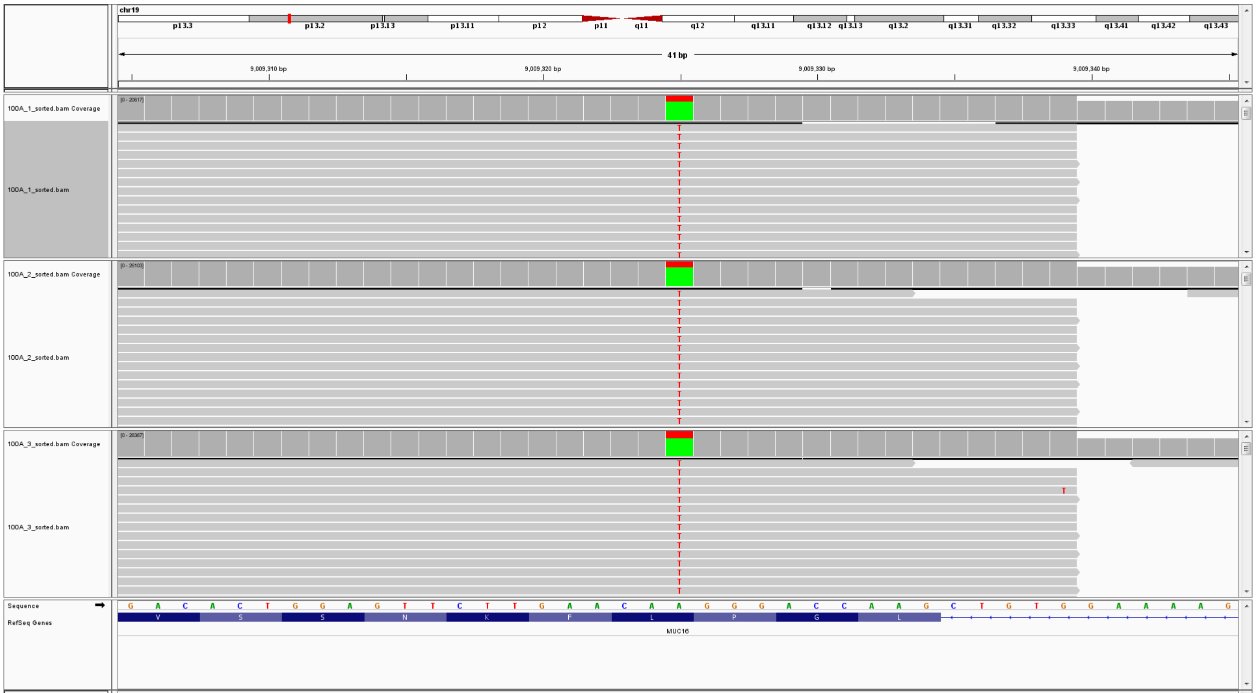
chr19
41 bp
9,009,310 bp
9,009,320 bp
9,009,330 bp
9,009,340 bp
100A_1_sorted.bam Coverage
100A_1_sorted.bam
100A_2_sorted.bam Coverage
100A_2_sorted.bam
100A_3_sorted.bam Coverage
100A_3_sorted.bam
Sequence
RefSeq Genes
MUC16

19

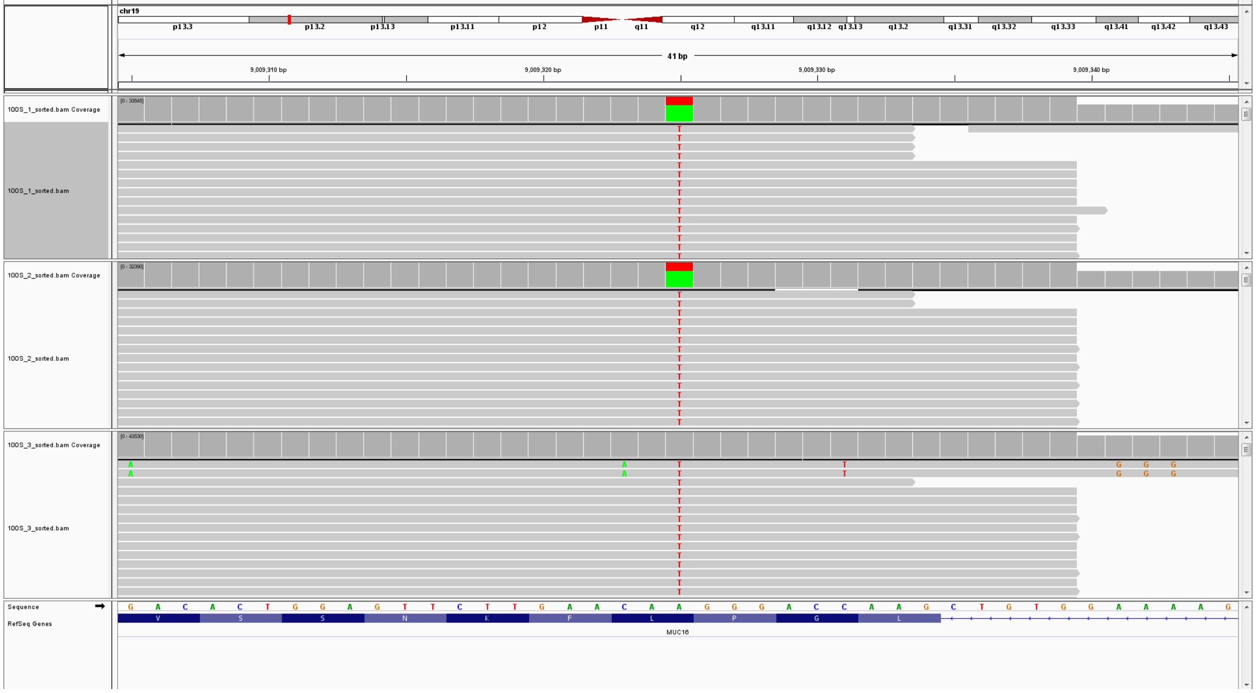
chr19
41 bp
9,009,310 bp
9,009,320 bp
9,009,330 bp
9,009,340 bp
100S_1_sorted.bam Coverage
100S_1_sorted.bam
100S_2_sorted.bam Coverage
100S_2_sorted.bam
100S_3_sorted.bam Coverage
100S_3_sorted.bam
Sequence
RefSeq Genes
MUC16

20

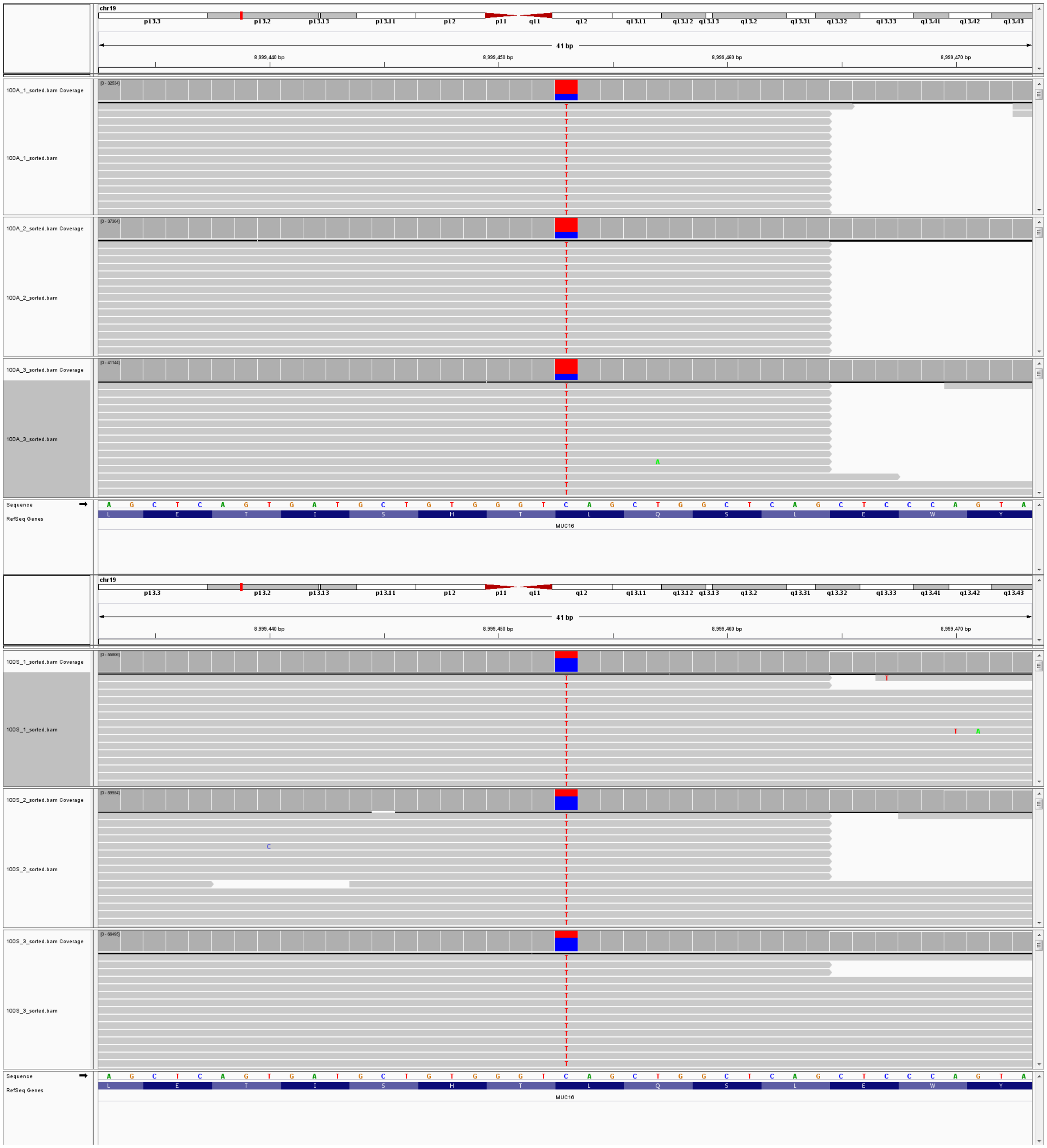


**Fig S3. SNVs with unbalanced alternative allele frequency.** Integrative Genomics Viewer [48] screen shot of three SNV examples - rs1800858 in RET, chr19:9009325 in MUC16 and rs11085765 in MUC16- shown for individual 1 (top) and individual 2 (botom). Three replicates for each individual are represented. The coverage track for each replicate shows a different color for the alternative and the reference allele frequency.

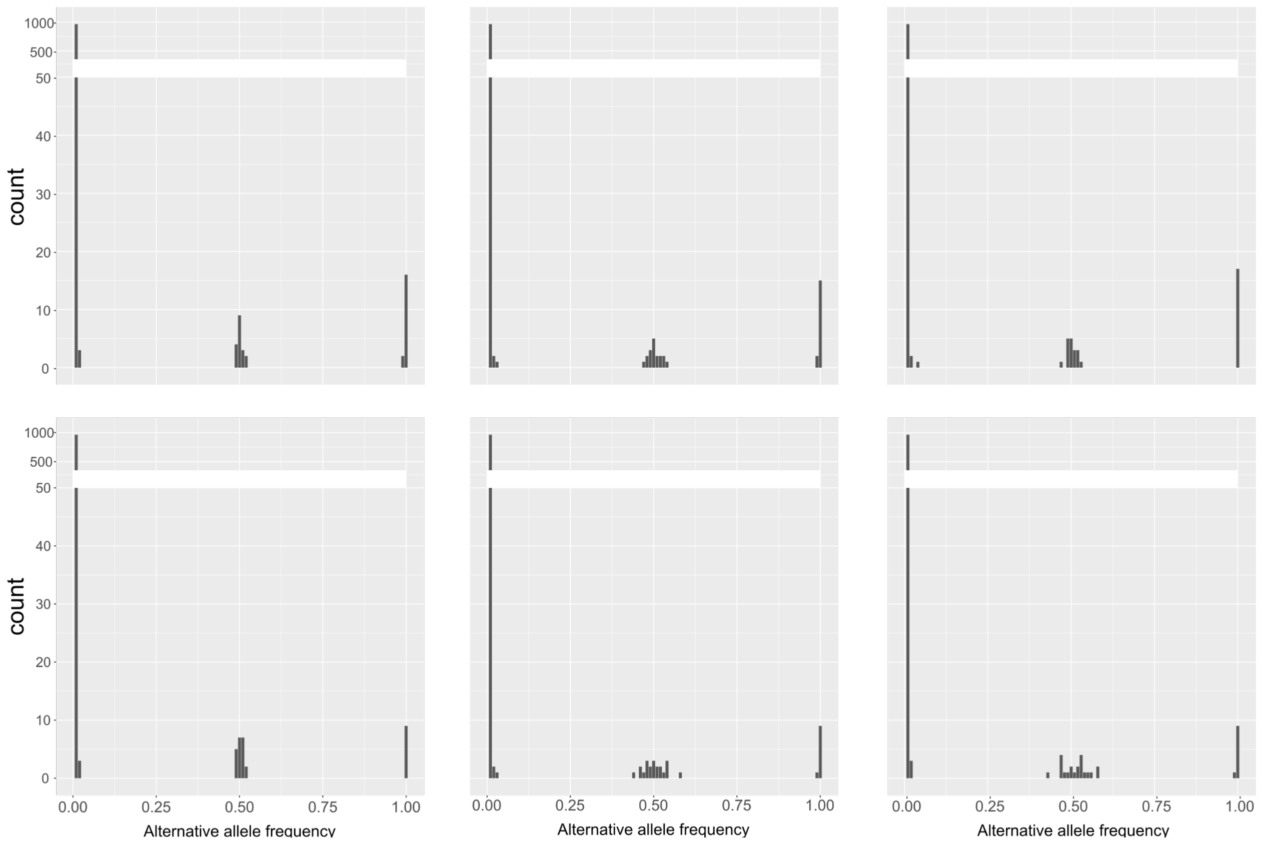


**Fig S4. Distribution of the alternative allele frequency after removing SNVs falling in primer sequence regions in targeted sequencing of urine DNA from two healthy individuals.** Three replicates for individual 1 (top) and for individual 2 (Bottom) were sequenced. The 1,000 most frequent SNVs in the ExAC project are presented here. Expected alternative allele frequency ~ 0, 0.5 and 1 for homozygous wild type, heterozygous and homozygous for the alternative allele, respectively.

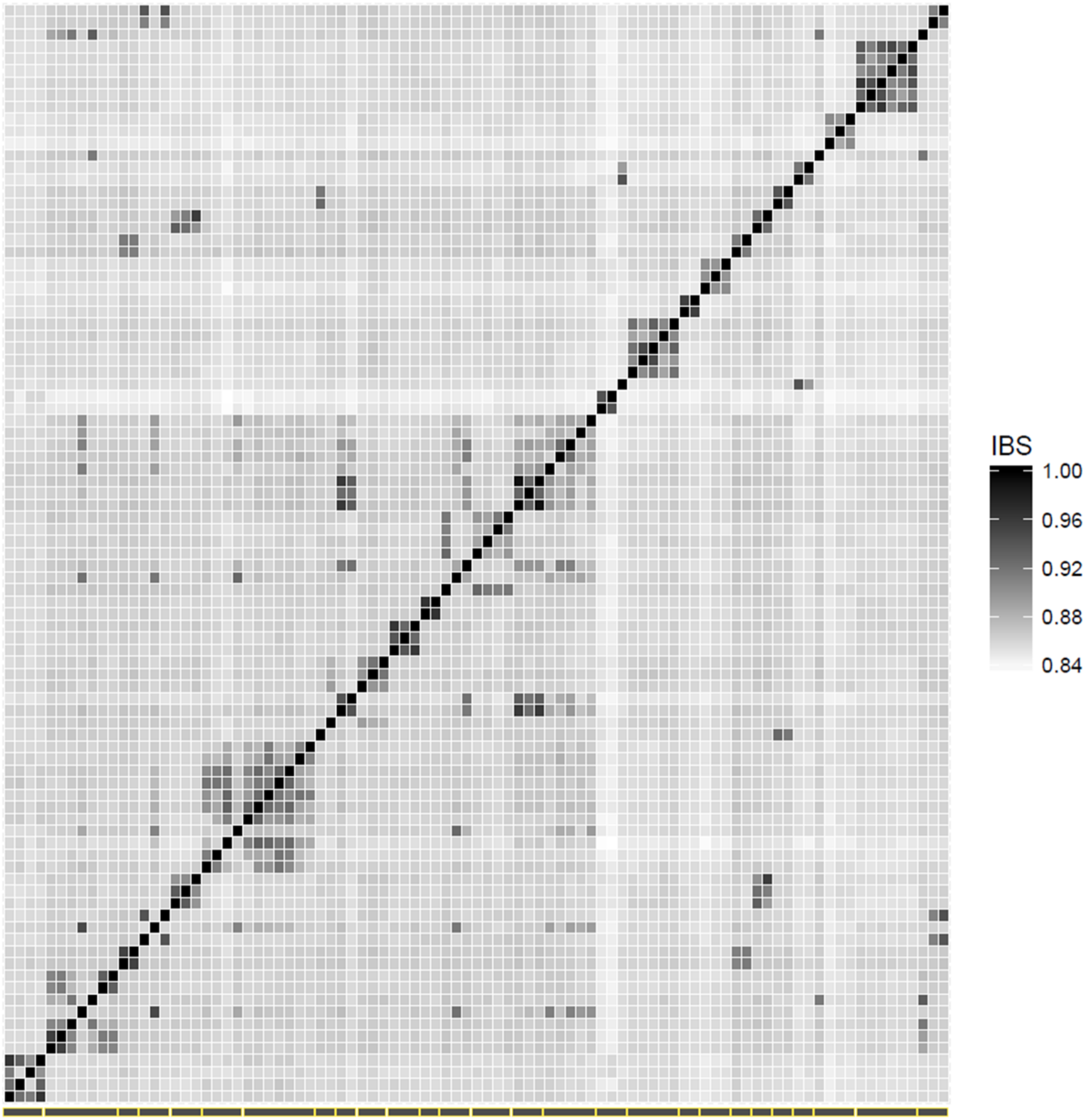


**S5 Fig. Heat map of 91 Qatari sibling whole genome sequences.** Different gradient indicates different identity by state values, as shown at the upper right corner. The black diagonal stands for the perfect relationship of each individual with himself. Twenty-seven nuclear families (grey boxes at the x axis) are represented on the bottom of the map.

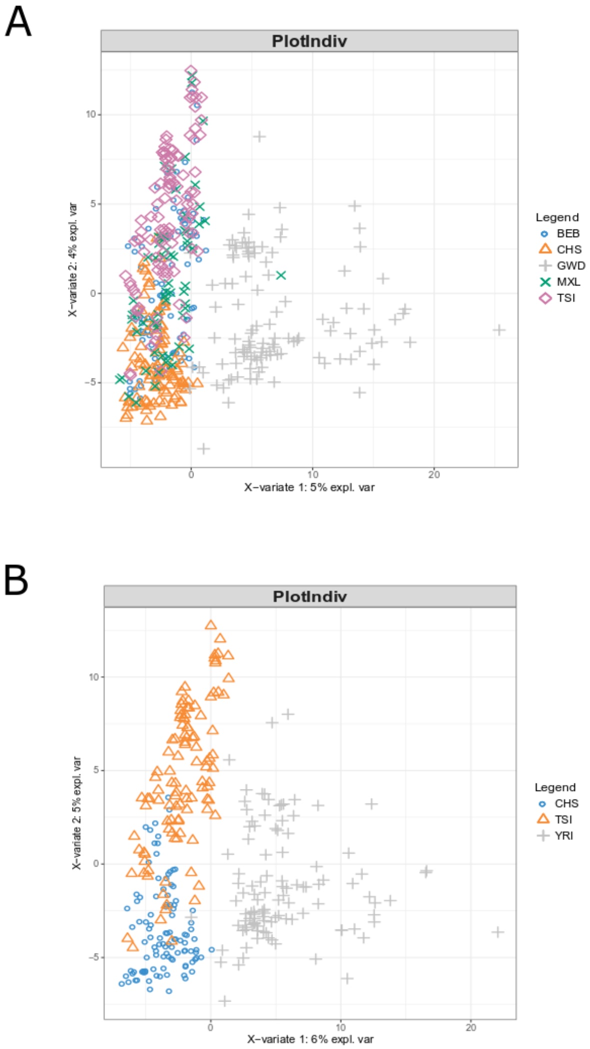


**S6 Fig. Partial least square analysis using A) five populations and B) three populations from the 1,000 genomes project.** Presented here the subpopulations with the highest cross validation accuracy: 54.8% for A) and 81.6% for B). A total of 1,000 SNVs were used in the analysis. BEB=Bengali from Bangladesh. CHS=Southern Han Chinese. GWD=Gambian in the Western Divisions in the Gambia. MXL=Mexican Ancestry from Los Angeles USA. TSI=Toscani in Italia. YRI=Yoruba in Ibadan in Nigeria.

**Table S1. Targeted genomic regions by the breast cancer DNA sequencing kit.**
